# Can non-disabled adults reproduce the stiff-knee gait of individuals with stroke?

**DOI:** 10.1101/2020.12.02.20242966

**Authors:** Odair Bacca, Melissa Celestino, José Barela, Anna Lima, Ana Barela

## Abstract

This study investigated whether a mechanical constraint of knee flexion in non-disabled individuals could help with reproducing the gait pattern of individuals with stroke. Eleven non-disabled adults (26.6±6.5 years old) and 12 individuals with stroke (52.0±12.8 years old) walked at a self-selected comfortable speed as kinematic and electromyographic data were acquired. Non-disabled adults also walked with an orthosis that limited to 45 degrees of knee flexion. The hip, knee, and ankle joint angles and the muscle activation of the rectus femoris, vastus medialis and lateralis, tibialis anterior, semitendinosus, biceps femoris, and gastrocnemius medialis and lateralis were analyzed. The results demonstrated that non-disabled adults presented similar lower limb excursion to individuals with stroke that affects most joints, although, they displayed a different muscle activation level for most muscles. These results suggest that a mechanical constraint of knee flexion leads to temporal and joint excursion alterations in the lower limb of non-disabled individuals, thereby enabling the reproduction of a gait pattern similar to individuals with stroke. It is also observed that these individuals use different strategies to control muscle activation, which might be related to the lack of control in coordinating muscle activation during gait that is present in individuals with stroke.

## Introduction

Limited knee flexion during the swing period of gait is a common movement disorder in individuals with stroke (Boudarhamet al., 2013; Olney, 1996). This is referred to as stiff-knee gait (Kerriganet al., 1991). This diminished knee flexion is generally associated with decreased gait speed, cadence, and stride length (Balaban and Tok, 2014; Balasubramanianet al., 2007; Chenet al., 2005) and the characteristic pattern inhibits toe clearance and leads to tripping, which in turn compromises stability and increases the risk of falls (Burpee and Lewek, 2015; Goldberget al., 2003; Merlo and Campanini, 2019).

Hyperactivity of the knee extensor muscles (Boudarhamet al., 2013; Rocheet al., 2015) and rectus femoris during the pre-swing phase and swing period of gait (Kerriganet al., 1991), and insufficient activation level of knee flexors (Flansbjeret al., 2006; Kligyteet al., 2003), including reduced activation of gastrocnemius muscles during pre-swing (Franciset al., 2013) are among the causes of stiff-knee gait in individuals with stroke. Although several studies have investigated the different aspects of gait in individuals with stroke, such as spatiotemporal parameters (Balaban and Tok, 2014; Titianovaet al., 2003), joint angles (Goldberget al., 2006; Woolley, 2001), and muscle activation (Den Otteret al., 2007; Lamontagneet al., 2000), the compensatory adaptations that might occur due to diminished knee flexion of the paretic limb still require investigation.

Individuals with stroke present with alterations in the lower limb joints during gait, and the knee joint, specifically, contributes markedly to mobility mainly during the swing period of gait (Lageet al., 1995). This joint is surrounded by several muscles, and each of them requires precise timing for activation and appropriate modulation of synergies during contraction (Shelburneet al., 2006). Cook et al. (1997) observed that constraining the knee flexion to 10 and 25 degrees decreased the ability to absorb impact during the duration of the gait of non-disabled young men. On the other hand, Lage et al. (1995) demonstrated that constraining knee flexion in non-disabled young adults to 0, 10, and 20 degrees increased the excursion of the ankle in dorsiflexion and hip flexion during the swing period of the constrained limb, as a compensatory adaptation. However, to our knowledge, only Hutin et al. (2011) compared the paretic knee of individuals with stroke to non-disabled adults walking with a constrained knee, exhibiting similar gait velocity, stride length, and thigh-shank coordination pattern during the swing period between both groups. Despite changes in the gait of non-disabled individuals due to mechanical constraints (Hutinet al., 2011), there are still certain issues that must be investigated, such as the excursion of lower limb joints other than the knee and the activation of the main muscles around the knee joint due to the diminished range of motion. Investigating the compensatory mechanisms related to knee constraints in non-disabled individuals would provide crucial insights regarding gait impairment and rehabilitation protocols for individuals with stroke.

The aim of this study was to investigate whether a mechanical constraint of knee flexion in non-disabled adults would lead to a gait pattern that is similar to that of individuals with stroke. More specifically, lower limb excursion and muscle activation was investigated for non-disabled adults walking with a constrained knee and individuals with chronic stroke walking freely. We hypothesized that non-disabled adults would present similar gait patterns of individuals with stroke on both the constrained/paretic and non-constrained/non-paretic limbs, increasing joint excursion of the hip and ankle joints, and activation of the rectus femoris and tibialis anterior muscles during the swing period of gait.

## Methods

### Sample

A total of 11 non-disabled males (control) and 12 individuals having experienced a stroke (7 women, 5 men), participated in this study. The inclusion criterion for the non-disabled adults was the absence of any known neurological or orthopedic impairment and musculoskeletal injuries that could compromise gait. The inclusion criteria for the individuals with stroke were more than 6 months past the stroke episode (either ischemic or hemorrhagic); ability to walk at least 10 m without assistance; ability to follow verbal commands; absence of any orthopedic or neurological impairment other than the stroke; and no chirurgical intervention, including botulinum toxin injection in the previous 6 months. Table 1 lists the general characteristics of both groups.

**Table 1.** General characteristics for non-disabled individuals (control) and individuals with stroke.

| Characteristic | Control | Stroke | p value |
| --- | --- | --- | --- |
| Age (yrs.)* | 26.6±6.5 | 52.0±12.8 | <0.001 |
| Mass (kg)* | 78.3±10.3 | 74.9±19.8 | 0.600 |
| Height (m)* | 1.77±0.06 | 1.64±0.09 | <0.001 |
| Body mass index (kg/m<sup>2</sup>)* | 24.9±2.9 | 27.4±5.0 | 0.145 |
| Male/Female | 11/0 | 5/7 | - |
| Time poststroke (mo)* | - | 102±72 | - |
| Ischemic/Hemorrhagic | - | 7/5 | - |
| Paretic/Constraint limb (Right/Left) | 11/0 | 5/7 | - |
| Fulg-Meyer score (max 84)* | - | ± | - |
| Functional independence score (max 126)* | - | ± | - |

\* Mean ± standard deviation values

The Institutional Review Board approved all the procedures, and all the participants signed a written consent form prior to the experimental session.

### Procedures

Reflective markers were placed bilaterally on the anterior and posterior superior iliac crest, lateral and medial femoral epicondyle, tibialis tuberosity, lateral and medial malleolus, inter malleolus, calcaneus, and first and fifth metatarsal heads. In addition, rigid clusters with non-collinear reflective markers were placed on the sacrum, thighs, and shanks. Passive disposable dual Ag/AgCl electrodes, with each circular conductive area having a 1-cm diameter and a 2-cm center-to-center distance, with double-differential preamplifiers (dual electrode, Noraxon, Inc.) were placed bilaterally on the belly of the following muscles: rectus femoris (RF), vastus medialis (VM), vastus lateralis (VL), tibialis anterior (TA), semitendinosus (ST), biceps femoris (BF), gastrocnemius medialis (GM), and gastrocnemius lateralis (GL). The skin area was shaved and cleansed with gauze soaked in alcohol, and electrodes were placed along the muscle fiber direction of each muscle following the guidelines of the Surface Electromyographic for Non-Invasive Assessment of Muscles (SENIAM) (Hermenset al., 2000). Body segments near the sensors and electrodes were slightly bandaged with elastic bands to avoid undesirable movement of sensors and their connectors.

All participants were made to walk, with their own walking shoes, at a self-selected comfortable speed on a 10-m walkway (“free walking condition”) equipped with 2 embedded force plates (Kistler, model 9286BA) covered with a thin rubber carpet. The non-disabled adults walked wearing an orthosis (Knee Ranger Lite-Endurance, Starben) that permitted full extension and limited flexion of their right knee up to 45 degrees (“constraint condition”). Prior to data acquisition, the participants practiced for a few trials and a calibration trial was acquired with all reflective markers placed on their body landmarks. After that, markers from the anterior superior iliac spine, posterior superior iliac spine, medial and lateral epicondyle of the femur, tibialis tuberosity, medial and lateral malleolus, and inter malleolus were removed, and a minimum of five trials were recorded for each participant and for the non-disabled adults for each experimental condition.

A computerized gait analysis system (VICON, Inc.) with 8 infrared cameras and a wireless electromiographic (EMG) system with 16 channels (Ultium EMG, Noraxon, Inc.) were used to acquire kinematic and EMG data. The EMG system had a gain of 500 and a common mode rejection ratio that was <-100dB, internal 24-bit sampling, input impedance that was >1000 MΩ, and a signal to noise ratio that was <1µV. All the data were acquired synchronously with the kinematic and force plate data at a frequency of 100 Hz and the EMG data was obtained at a frequency of 2000 Hz.

### Data analysis

Two consecutive and steady state strides (right and left for non-disabled adults, and paretic and non-paretic for individuals with stroke) per trial by each participant were analyzed for a total of 5 trials for each experimental condition. Foot contacts and toe-offs during each stride were identified using force plate data and the foot vertical velocity (O’connoret al., 2007) was considered for the subsequent calculation of the pre-swing phase and swing period (Perry, 1992).

The hip, knee, and ankle joint angles were calculated with The MotionMonitor (Innovative Sports Training, Inc.), and subsequent analyses were performed using custom routines written in MATLAB (Mathworks, Inc.). All the data were digitally filtered using a 4th order and zero-lag Butterworth filter. Low pass filtering at 6 Hz was performed on the kinematic data and band-pass filtering at 20–400 Hz was conducted on the EMG data, which was subsequently full-wave rectified and low-pass filtered at 5 Hz to obtain the linear envelope. The amplitude of the EMG data was obtained by calculating the root mean square (RMS) of the signal over a 50-ms window with a 20-ms overlap (Bonatoet al., 1998). This was calculated for the pre-swing phase and swing period of gait. Data for each muscle were normalized by the maximum value obtained across all free-walking trials (Burdenet al., 2003). All gait cycles were normalized in time from 0% to 100%, and these cycles were averaged to obtain the mean cycle for each participant. This procedure was repeated to determine the mean cycle among participants in each group.

The variables used in this study were: mean walking speed, calculated as the ratio between the distance covered by two strides and its duration (determined by the position of the sacrum marker); duration of the pre-swing phase and swing period; hip, knee, and ankle joint angles at toe-off and maximum flexion (hip and knee) and dorsiflexion (ankle) during the swing period; and RMS of RF, VM, VL, TA, ST, BF, GM, and GL muscles during the pre-swing phase and swing period. Since the hip and ankle angles present negative values of extension and plantar flexion, respectively, we calculated the absolute angles for all joints for the statistical analyses, with 180 degrees for the hip and knee and 90 degrees for the ankle equivalent to neutral position.

### Statistical analyses

An analysis of variance (ANOVA) was initially employed to compare the mean walking speed of both groups (non-disabled adults walking with a constrained knee and individuals with stroke walking freely). Since there was a group difference, the mean walking speed was employed as a covariate. Multivariate analyses of covariance (MANCOVA), having as factors group and limb (constrained/paretic, non-constrained/non-paretic), the last one considered as repeated-measure, were employed. The dependent variables were the pre-swing and swing durations for the first MANCOVA; hip, knee, and ankle joint angles at toe-off for the second MANCOVA; maximum hip, knee, and ankle joint angles during swing period for the third MANCOVA; RMS of the following muscle groups: RF, VM, and VL; ST and BF; GM and GL, during pre-swing phase and swing period, respectively, for the remaining MANCOVAs. Two ANCOVAs with the same factors were employed for the RMS of the TA muscle during the pre-swing phase and swing period, respectively. When necessary, univariate analyses and Tukey’s test for post hoc analysis were employed. An alpha level of 0.05 was set for all the statistical tests, which were performed using SPSS software.

## Results

Table 2 presents the mean (±SE) values of mean walking speed, pre-swing phase, and swing period durations. The ANOVA for mean walking speed revealed that non-disabled individuals with a constrained limb walked faster than individuals with stroke (F_1,21_=26.73, p<0.001). The MANCOVA for pre-swing and swing durations revealed a limb effect (Wilks’ Lambda=0.35, F_2,19_=17.69, p<0.001). Univariate analyses indicated longer pre-swing (F_1,20_=9.26, p=0.006) and swing (F_1,20_=7.64, p=0.012) durations in the constrained and paretic limbs compared to that of non-constrained and non-paretic limbs.

**Table 2.** Mean (± SD) values of mean walking speed, pre-swing and swing durations of non-disabled individuals (control) and individuals with stroke.

| Variable | Control |  | Stroke |  |
| --- | --- | --- | --- | --- |
|  | Constrained | Non-constrained | Paretic | Non-paretic |
| Walking speed (m/s) | 1.25±0.06 | | 0.79±0.06 | |
| Pre-swing duration (%) | 10.2±1.7 | 11.1±0.9 | 19.9±1.6 | 14.8±0.9 |
| Swing duration (%) | 40.1±1.4 | 38.6±1.3 | 35.9±1.3 | 29.4±1.3 |

Figure 1 displays the mean (±SD) gait cycle of the hip, knee, and ankle joint angle. Overall, both groups presented roughly similar patterns, with reduced knee flexion of constrained and paretic limbs during the swing period. The hip (pre-swing) and ankle (swing) joints of the paretic limb seem to remain closer to neutral position than those of the constrained, non-constrained, and non-paretic limbs.

**Figure 1.**
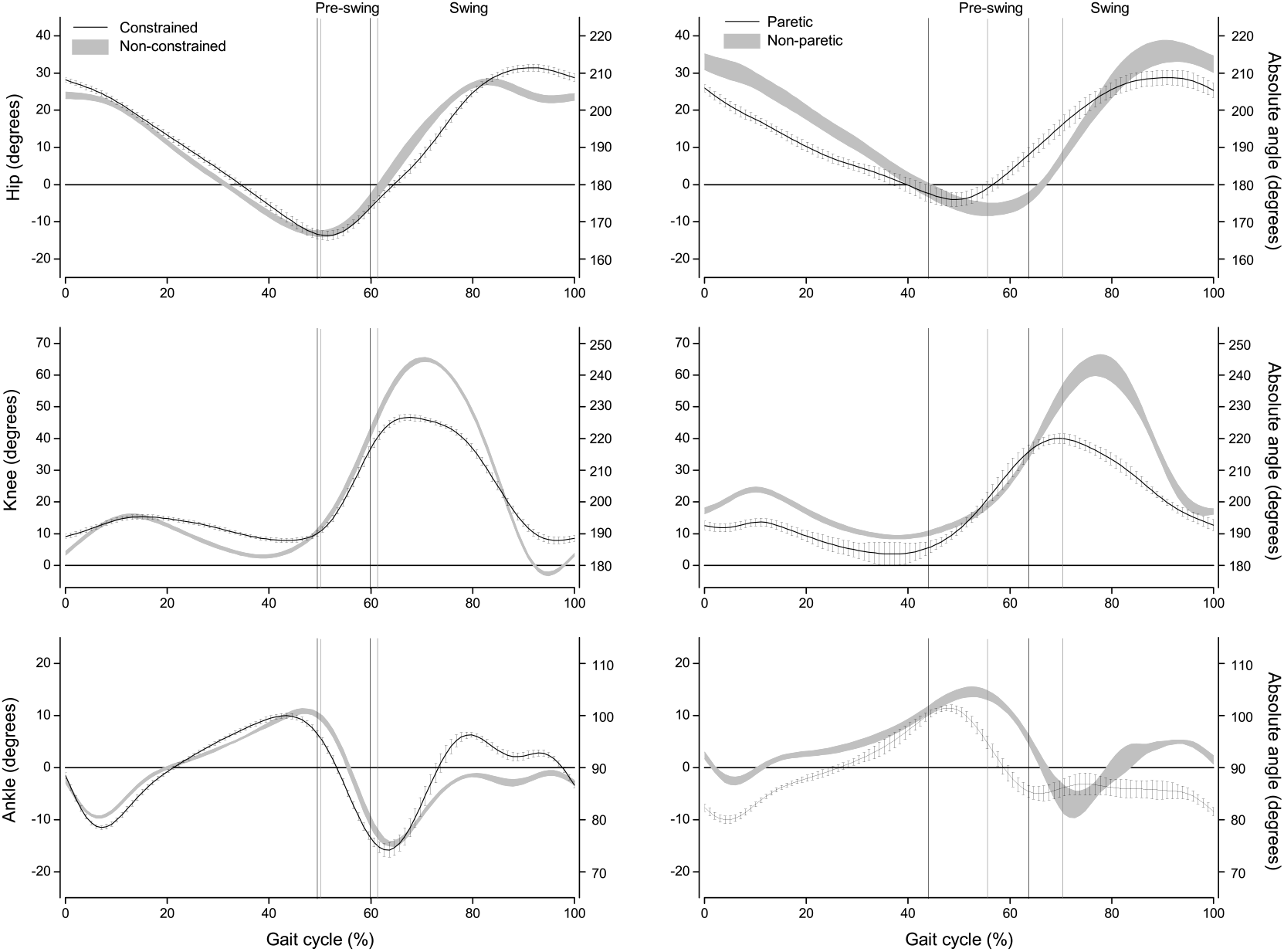
Mean (±SD) gait cycle of hip, knee, and ankle joint angles for the non-disabled adults walking with a constrained knee flexion movement and individuals with stroke walking freely. Note: constrained/paretic limbs are represented by a line and non-constrained/non-paretic limbs are represented by the grey area.

Figure 2 depicts the mean (±SE) values of joint angles at toe-off (A) and during the swing period (B). The MANCOVA for joint angles at toe-off revealed limb effect (Wilks’ Lambda=0.44, F_3,18_=6.67, p=0.002) and group and limb interaction (Wilks’ Lambda=0.56, F_3,18_=4.71, p=0.013). Univariate analyses indicated limb effect for the knee (F_1,20_=10.77, p=0.004) and ankle (F_1,20_=6.94, p=0.016) joints, and group and limb interaction for the ankle joint (F_1,20_=11.54, p=0.003). At toe-off, the knee of the non-constrained and non-paretic limbs presented more flexion than the constrained and paretic limbs, and the ankle of the constraint limb presented less dorsiflexion than that of the paretic limb.

**Figure 2.**
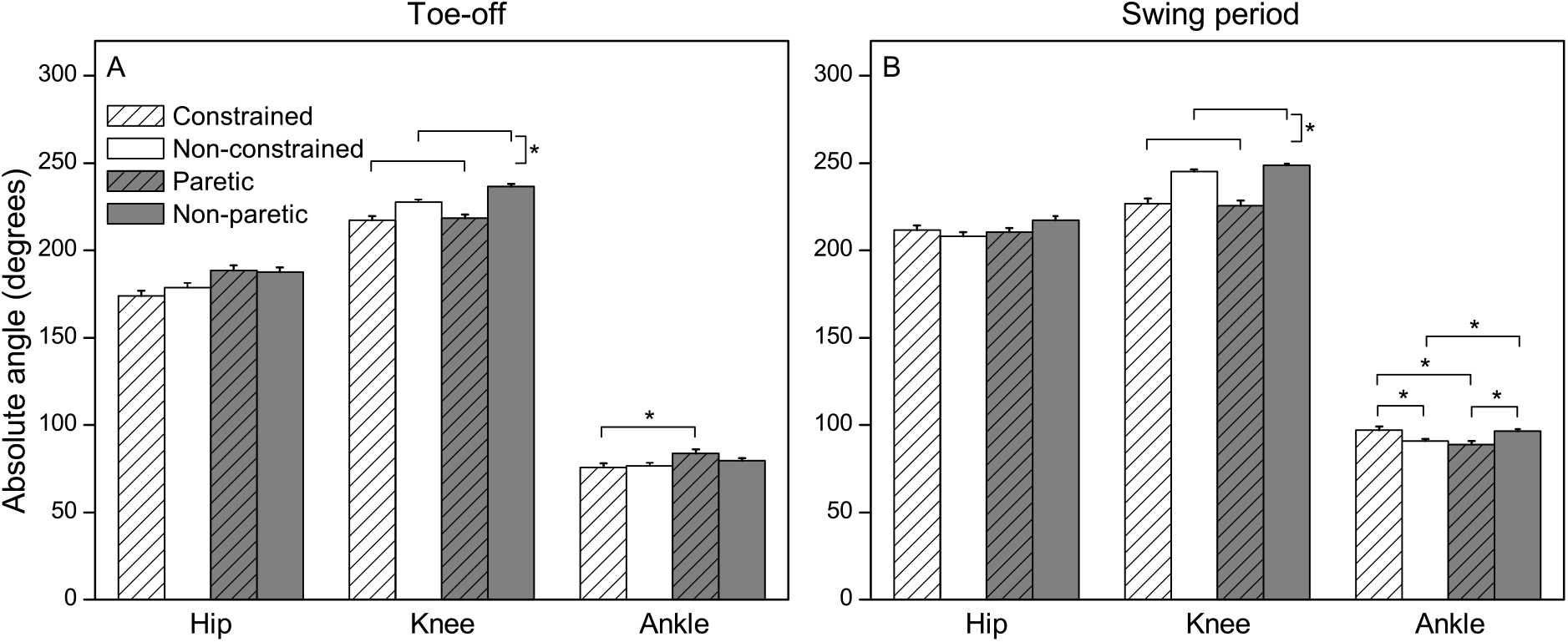
Mean (±SE) values of hip, knee, and ankle absolute angles at toe-off (A) and during swing period (B) for the non-disabled adults walking with a constrained knee flexion movement and individuals with stroke walking freely.

The MANCOVA for the maximum angles during the swing period revealed limb effect (Wilks’ Lambda=0.61, F_3,18_=3.88, p=0.027), and group and limb interaction (Wilks’ Lambda=0.64, F_3,18_=3.37, p=0.041). Univariate analyses indicated limb effect for the knee (F_1,20_=9.73, p=0.005) and ankle (F_1,20_=4.49, p=0.047) joints, and group and limb interaction for the ankle joint (F_1,20_=4.48, p=0.047). The knee of the non-constrained and non-paretic limbs presented more flexion than that of the constrained and paretic limbs. The ankle of the constrained limb exhibited more dorsiflexion than that of the non-constrained and paretic limbs; the ankle of the paretic limb presented less dorsiflexion than that of the non-paretic limb, and the ankle of the non-constrained limb exhibited lesser dorsiflexion than that of the non-paretic limb.

Figures 3 and 4 depict the mean (±SD) gait cycle of muscle activation of non-disabled adults and individuals with stroke. Overall, it seems that the activation patterns of the RF, VM, VL, and TA muscles (Fig. 3) of non-disabled adults are similar in both constrained and non-constrained limbs with distinct activation peaks and silent periods. The activation pattern of the individuals with stroke in these muscles is roughly similar for paretic and non-paretic limbs, except for the TA muscle during the pre-swing phase, which seems more active in the paretic limb. Individuals with stroke seem to maintain a level of activation for these muscles throughout the gait cycle, in contrast to the non-disabled adults.

**Figure 3.**
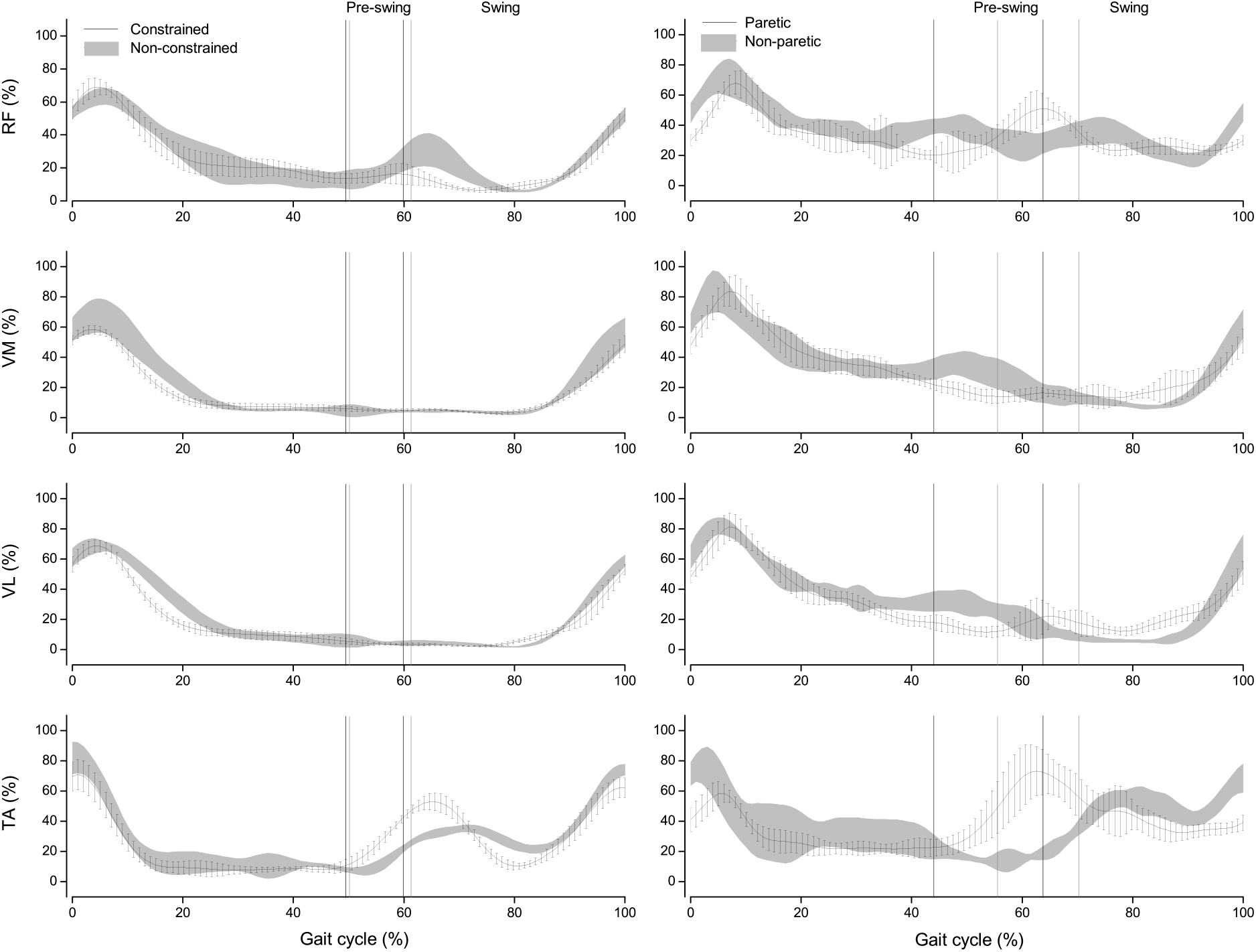
Mean (±SD) gait cycle of rectus femoris (RF), vastus medialis (VM), vastus lateralis (VL), and tibialis anterior (TA) muscles for the non-disabled adults walking with a constrained knee flexion movement and individuals with stroke walking freely. Note: constrained/paretic limbs are represented by a line and non-constrained/non-paretic limbs are represented by the grey area.

**Figure 4.**
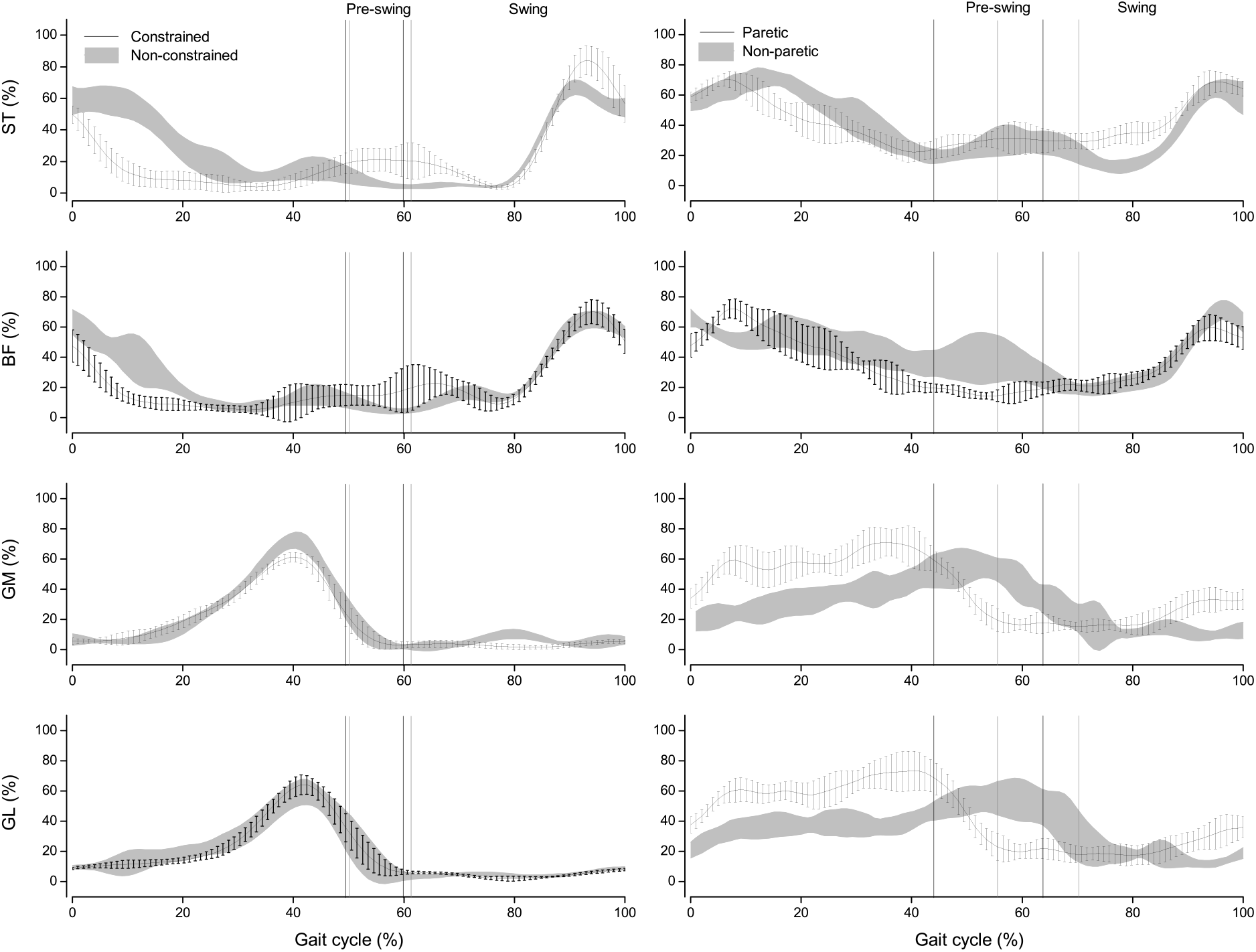
Mean (±SD) gait cycle of semitendinosus (ST), biceps femoris (BF), gastrocnemius medialis (GM), and gastrocnemius lateralis (GL) muscles for the non-disabled adults walking with a constrained knee flexion movement and individuals with stroke walking freely. Note: constrained/paretic limbs are represented by a line and non-constrained/non-paretic limbs are represented by the grey area.

In terms of ST, BF, GM, and GL muscles (Fig. 4), the non-disabled adults presented similar activation patterns for both constrained and non-constrained limbs for the GM and GL muscles, and slightly different activation patterns at the beginning of the gait cycle for the ST and BF muscles. The individuals with stroke presented roughly similar activation patterns between paretic and non-paretic limbs for ST and BF muscles, as GM and GL in the paretic limb were more active prior to the pre-swing phase and seemed more silent afterwards. In the non-paretic limb, they were active until the beginning of the swing period. Individuals with stroke seem to maintain a level of activation for the ST, BF, GM, and GL muscles throughout the gait cycle.

Figure 5 depicts the mean (±SE) values of the RMS of selected muscles during the pre-swing phase and swing period. The MANCOVA for the RF, VM, and VL muscles during the pre-swing phase revealed a group effect (Wilks’ Lambda=0.65, F_3,18_=3.29, p=0.045) and group and limb interaction (Wilks’ Lambda=0.62, F_3,18_=3.75, p=0.030). Univariate analyses indicated a group effect for the VM (F_1,20_=6.05, p=0.023) and VL (F_1,20_=9.77, p=0.005) muscles, and group and limb interaction for the RF muscle (F_1,20_=4.49, p=0.030). Non-disabled adults presented a lower RMS than individuals with stroke for the VM and VL muscles, and the constrained limb presented lower RMS for the RF muscle than that of the paretic limb.

**Figure 5.**
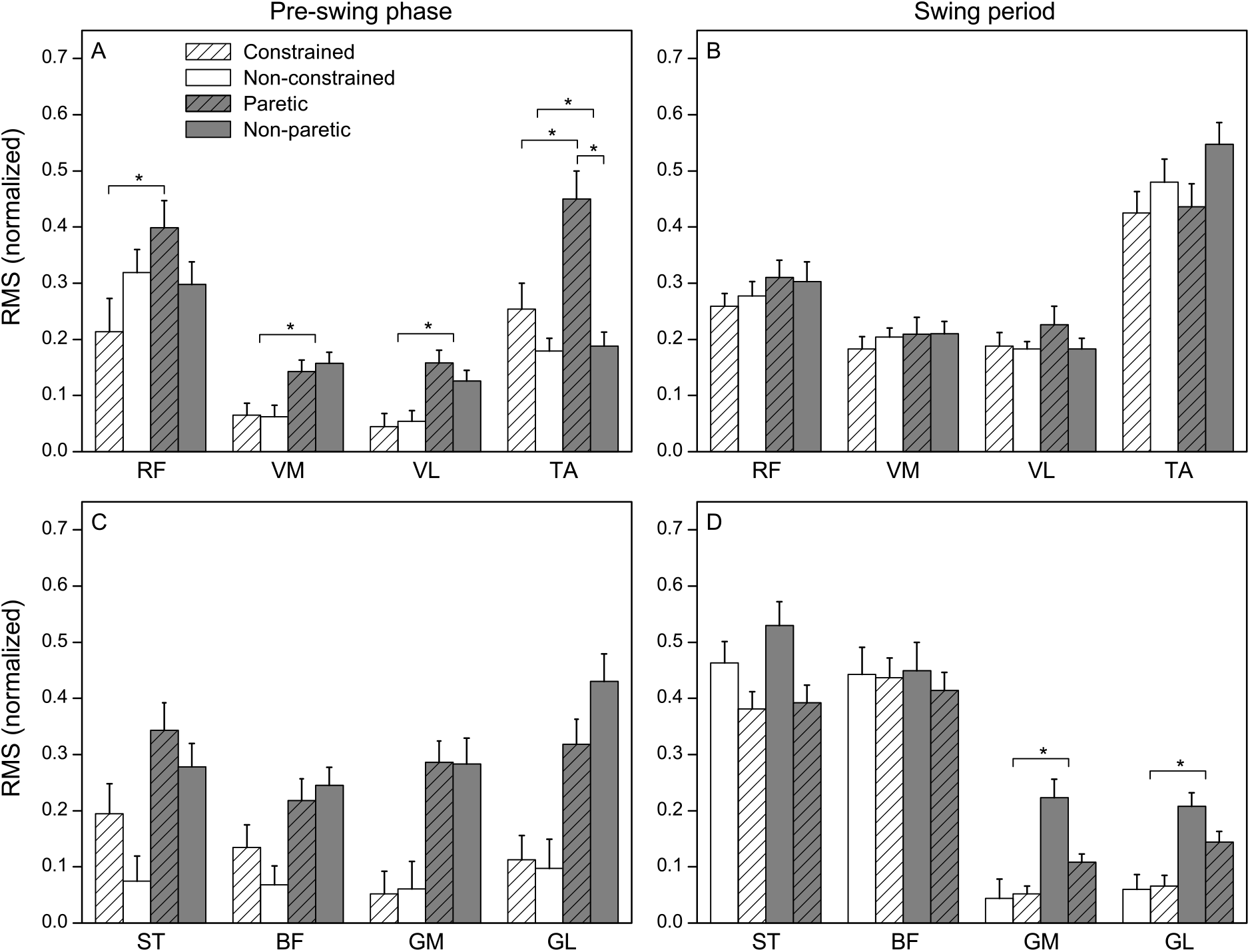
Mean (±SE) values of rectus femoris (RF), vastus medialis (VM), vastus lateralis (VL), tibialis anterior (TA), semitendinosus (ST), biceps femoris (BF), gastrocnemius medialis (GM), and gastrocnemius lateralis (GL) during pre-swing phase (A, C) and swing period (B, D) for the non-disabled adults walking with a constrained knee flexion movement and individuals with stroke walking freely.

The ANCOVA for the TA muscle during the pre-swing phase revealed a group effect (F_1,20_=6.57, p=0.019) and group and limb interaction (F_1,20_=11.42, p=0.003). Non-disabled adults presented a RMS that was lower than that for individuals with stroke, and the constrained and non-paretic limbs presented a lower RMS than the paretic limb.

The MANCOVA for the GM and GL muscles during the swing period revealed a group effect only (Wilks’ Lambda=0.71, F_2,19_=3.95, p=0.037). Univariate analyses indicated a group effect for both GM (F_1,20_=4.84, p=0.040) and GL (F_1,20_=7.22, p=0.014) muscles. Non-disabled adults presented a lower RMS than individuals with stroke for both muscles. No other differences were found for the remaining muscles during either the pre-swing phase or swing period.

## Discussion

We investigated whether a mechanical constraint of knee flexion in non-disabled adults would reproduce the gait pattern, in terms of joint excursion and muscle activation of the lower limbs, of individuals with stroke. In general, the results indicated that non-disabled adults walking with a constrained limb that limited knee flexion presented similar lower limb excursion to individuals with stroke in most joints and similar temporal organization; though, with a different muscle activation (pattern and level) for hip, knee, and ankle movements. Therefore, these results partially confirmed our hypotheses.

Constrained and paretic limbs presented longer swing durations, which is a common strategy adopted by individuals with stroke (Chenet al., 2005; Titianovaet al., 2003) in an attempt to decrease the time that the body must support itself while loaded on the paretic limb (Balaban and Tok, 2014; Hsuet al., 2003; Titianovaet al., 2003). This is most likely due to the difficulty in moving the paretic limb forward (Chen et al., 2003; Neptuneet al., 2004). It is remarkable that a simple mechanical restriction to the knee joint excursion could also lead to such an asymmetrical temporal organization of the gait cycle in non-disabled adults, as revealed by our results.

A mechanical constraint also caused critical changes in joint excursion in non-disabled adults. The most prominent change was, as expected, in the knee of the constrained limb, mimicking the paretic limb, which might be a signature of stiff-knee gait in individuals with stroke (Campaniniet al., 2013; Goldberget al., 2004; Olneyet al., 1991). The ankle joint was also affected, leading to opposite alterations observed in individuals with stroke. Usually, individuals with stroke do not present dorsiflexion in the paretic limb during the swing period, and non-disabled individuals increase dorsiflexion in the constrained limb. This finding can be attributed to the different selection of muscle activation, as individuals with stroke present lack of control to activate plantar flexor muscles in the paretic limb during the pre-swing phase (Franciset al., 2013; Wanget al., 2017). Therefore, we can suggest that a mechanical constraint in a knee joint imposes similar restrictions on the non-disabled individuals at the knee joint motion, impacting their gait pattern, but leading to different kinematic solutions in the adjacent lower limb joints, which does not seem to be available to individuals with stroke.

The absence or lack of possible solutions observed in individuals with stroke is due to difficulties in selectively activating the lower limb muscles (Den Otteret al., 2007; Srivastavaet al., 2019). These individuals presented higher activation of the rectus femoris, vastus medialis, and vastus lateralis muscles during pre-swing as compared to the constrained limb. The hyperactivity of the rectus femoris muscle presented by individuals with stroke corroborates the findings of previous studies (Boudarhamet al., 2013; Grosset al., 2017; Rocheet al., 2015), indicating that it contributes to diminished knee flexion (Piazza and Delp, 1996; Reinboltet al., 2008). The higher activation of the vastus medialis and lateralis muscles during pre-swing is associated with insufficient velocity of knee flexion at toe-off, which in turn contributes to diminished knee flexion during the swing period (Goldberget al., 2004). The lack of similarities in muscle activation between non-disabled individuals and those with stroke might be attributed to the more efficient strategies presented by non-disabled individuals, since they do not have difficulty dissociating the activation of agonist and antagonist muscles around the knee joint.

In conclusion, a mechanical constraint was devised that could flex the knee, such that the temporal and joint excursion alterations in the lower limb of non-disabled adults simulated a gait pattern similar to that of individuals with stroke. However, non-disabled individuals presented different strategies to control muscle activation, which might be related to a lack of control in individuals with stroke in coordinating muscle activation during gait.

## Data Availability

All data will be available upon request

